# Cortical thickness and grey-matter volume anomaly detection in individual MRI scans: Comparison of two methods

**DOI:** 10.1101/2024.02.27.24303078

**Authors:** David Romascano, Michael Rebsamen, Piotr Radojewski, Timo Blattner, Richard McKinley, Roland Wiest, Christian Rummel

## Abstract

Over the past decades, morphometric analysis of brain MRI has contributed substantially to the understanding of healthy brain structure, development and aging as well as to improved characterisation of disease related pathologies. Certified commercial tools based on normative modeling of these metrics are meanwhile available for diagnostic purposes, but they are cost intensive and their clinical evaluation is still in its infancy. Here we have compared the performance of “ScanOMetrics”, an open-source research-level tool for detection of statistical anomalies in individual MRI scans, depending on whether it is operated on the output of FreeSurfer or of the deep learning based brain morphometry tool DL+DiReCT. When applied to the public OASIS3 dataset, containing patients with Alzheimer’s disease (AD) and healthy controls (HC), cortical thickness anomalies in patient scans were mainly detected in regions that are known as predilection areas of cortical atrophy in AD, regardless of the software used for extraction of the metrics. By contrast, anomaly detections in HCs were up to twenty-fold reduced and spatially unspecific using both DL+DiReCT and FreeSurfer. Progression of the atrophy pattern with clinical dementia rating (CDR) was clearly observable with both methods. DL+DiReCT provided results in less than 25 minutes, more than 15 times faster than FreeSurfer. This difference in computation time might be relevant when considering application of this or similar methodology as diagnostic decision support for neuroradiologists.

## Introduction

Many pathological processes affecting the central nervous system (CNS) have an impact on its structural organization. Various forms of brain morphometry have made it possible to describe brain shape mathematically, yielding variables for statistical evaluation, which have made important contributions towards a better understanding of healthy brain development and aging as well as to disease manifestation and mechanisms (see e.g. Mills et al., 2021; Statsenko et al., 2022; McCutcheon et al., 2023 for recent examples). Large group studies have demonstrated that metrics derived from routine structural MRI scans are sensitive to pathological brain changes (see e.g. Whelan et al., 2018; Laansma et al., 2021). For this reason, brain morphometric variables have been included as outcome measures in recent clinical trials (e.g. National Library of Medicine [NLM], NCT04860947 for the prediction of disease progression in multiple sclerosis, or NLM NCT06155942 for the use of morphometry as a biomarker for Parkinson’s disease).

Surface based analysis (SBA) is a variant of brain morphometry, that attempts to represent the two-dimensional geometry of the cortex by tesselating the interface between white matter (WM) and gray matter (GM) with a mesh and estimating region specific metrics like the GM volume (GMV), cortical surface area (CSA) or cortical thickness (CTh). During the last two decades, substantial efforts have been invested into providing software to extract precise and accurate SBA metrics from MRI scans. For research purposes, FreeSurfer (Dale et al., 1999; Fischl et al., 1999a, 1999b; Fischl & Dale, 2000) has become the most widely used automated tool. Among its advantages are its free availability and extremely high acceptance and understanding by the community, which has led to more than 2’800 scientific publications (PubMed search on 2024/02/17).

Sensitivity of SBA metrics to pathological processes has been mostly established through cross-sectional and longitudinal *group studies* (see e.g. de Figueiredo et al., 2021; Alkal et al., 2021; Nkrumah et al., 2023; Fortea et al., 2023 for recent examples). In contrast, normative modeling aims at a quantitative evaluation of *single subject* scans by establishing healthy developmental trajectories and prediction intervals of SBA metrics in a reference population. It is a powerful tool to detect statistical anomalies at the individual level, making it much better suited to support personalized diagnostics and decision making (Marquand et al., 2016; Potvin et al., 2017; Marquand et al., 2019; Potvin et al., 2021; Ge et al., 2023). In the meantime, CE-marked and FDA-approved commercial tools for clinical decision support by brain morphometry and normative modeling have become available for application in various forms of dementia (Pemberton et al., 2021) and in patients with MS (Mendelson et al., 2023).

To provide reliable predictions, the models should be derived from large normative databases (Rutherford et al., 2022). In the field of MRI, suitable datasets have recently become available as public resources and open doors towards the application of normative models in clinical settings. Since MRI acquisition settings like scanner type (Sinnecker et al., 2022) or scanning protocol (Rebsamen et al., 2023b) have been demonstrated to influence SBA estimates, control for these confounders by harmonization procedures is required (see e.g. Fortin et al., 2018). Our own work in the direction of normative modeling has demonstrated screening test characteristics of automated regional SBA metrics in patients with temporal lobe epilepsy (i.e. large negative predictive values, while positive predictive values were only moderate; Rummel et al., 2017) and provided markers for regional atrophy progression in patients with multiple sclerosis (Rummel et al., 2018).

One of the remaining obstacles hindering the use of SBA normative modeling as a decision support tool in the clinical routine is the long computation time required for tools like FreeSurfer to process a single MRI scan, which is in the order of ten hours on the central processing unit (CPU) of a current standard desktop computer. Indeed, to practically contribute information to clinical diagnostics, processing times should ideally be reduced to the order of minutes, to enable patient evaluation on demand or at least within the same shift. To overcome this limitation, new tools leveraging deep learning (DL) and convolutional neural networks (CNN) running on graphical processing units (GPU) have become available for SBA, like for example FastSurfer (Henschel et al., 2020). DL+DiReCT (Rebsamen et al., 2020, 2023a) and CortexMorph (McKinley & Rummel, 2023) are alternative approaches to DL-based estimation of CTh. A recent comparative study revealed that not only did DL+DiReCT substantially outperform FreeSurfer in terms of computation time required to estimate CTh, but it also provided comparable scan-rescan reproducibility and estimated atrophy rates (Rebsamen et al., 2020). Importantly, DL+DiReCT was shown superior to FreeSurfer (both cross-sectional and longitudinal) in terms of sensitivity to simulated cortical thinning, especially when the introduced atrophy was weak (Rusak et al., 2022).

The purpose of this work was to explore the performance of our normative modeling approach (“ScanOMetrics”, Rummel et al., 2017, 2018) on metrics derived from DL+DiReCT and FreeSurfer. To achieve full reproducibility of our results, we focussed the analysis on OASIS3 (Open Access Series of Imaging Studies; LaMontagne et al., 2019), a large and freely available dataset containing clinical grade high-resolution isotropic T1-weighted MRI scans of patients with Alzheimer’s disease (AD) and healthy controls (HC). We restricted our software comparison to the jointly available SBA metrics of the Desikan-Killiany atlas (Desikan et al., 2006), namely regional cortical GMV as well as regional mean and standard deviation of the CTh. The ability to detect regional outliers was compared between the two processing tools and the accumulation of anomalies in brain regions that are known for atrophy in AD group studies was assessed.

Our hypotheses were the following: Based on results by Rusak et al. (2022) and ourselves (Rebsamen et al. 2020), we expected normative models within ScanOMetrics to provide (1) more narrow distributions of fit residues, (2) higher scan-rescan reproducibility, as well as (3) more pronounced and more specific atrophy patterns in patients when using DL+DiReCT instead of FreeSurfer metrics. Based on previous work using PET and MRI imaging (Jansen et al., 2022; Verdi et al., 2023), we expected that (4) the AD group would yield a higher percentage of individual scans labeled as anomalous than a leave-one-out cross-validation (LOOCV) in the HC group. Finally, we hypothesized that (5) normative modeling at the level of individual scans/patients shows heterogeneous anomaly patterns. When averaging the individual anomaly maps over the whole group, the shared anomaly motifs should, however, be similar to the map obtained when testing for statistical differences between the entire AD and HC groups (i.e. effect sizes in a group analysis).

## Materials and methods

All software tools used in this paper are open-source. The Python3 implementation of ScanOMetrics is available at https://github.com/SCAN-NRAD/ScanOMetrics. A code description is given in the Supplementary Materials and a more detailed documentation with tutorial is available at https://scanometrics.readthedocs.io. FreeSurfer can be downloaded from https://surfer.nmr.mgh.harvard.edu/ and DL+DiReCT is available at https://github.com/SCAN-NRAD/DL-DiReCT.

### OASIS3 dataset

The OASIS3 dataset (Open Access Series of Imaging Studies, LaMontagne et al., 2019) is publicly available at www.oasis-brains.org/ and contains NIFTI files of 2’643 high-resolution (voxel sizes in the order of 1 mm x 1 mm x 1 mm) isotropic T1-weighted MRI scans from 1’038 participants. We report specific scan IDs throughout the manuscript to illustrate results and discussion topics. Such IDs cannot be used to identify subjects, as the OASIS3 website states that “all participants were assigned a new random identifier”. All scans were acquired at two field strengths using Siemens MRI scanners: Magnetom Sonata and Avanto (1.5T, 42 scans) as well as Biograph mMR and Magnetom Trio (both 3T, 2’601 scans). 2’014 scans are from subjects considered HCs with normal cognition (clinical dementia rating CDR=0), 420 scans are from undetermined cases with CDR=0.5, and 209 scans correspond to patients with established AD having CDR >= 1, leading to 629 scans with CDR > 0.

Of the 2’014 HC scans, 87 are from 41 subjects that had a mixture of scans with CDR=0 and CDR>=0.5 (“converters” between normal cognition and suspicion or established impairment). Those scans were excluded from building our normative model, which was therefore based on 1’927 scans from 696 non-converting subjects, see Table 1 for demographic information. The 41 ‘converter’ subjects were instead used to investigate the change trajectories between the first and follow-up scans longitudinally.

**Table 1:** Demographic characteristics of the used OASIS3 subgroups. To increase clarity, 60 scans (2.2%) from 15 patients with CDR changing between different levels of CDR>=0.5 are not included here.

|  |  |  |  |  |
| --- | --- | --- | --- | --- |
| | CDR=0,<br>'non-converters'<br>used for<br>normative<br>models | subjects with<br>CDR=0 and<br>$\geq 0.5$,<br>'converters'<br>used for<br>follow-up | CDR=0.5,<br>'undetermined'<br>cases | CDR$\geq 1$,<br>'established'<br>dementia |

Table 1: Demographic characteristics of the used OASIS3 subgroups. To increase clarity, 60 scans (2.2%) from 15 patients with CDR changing between different levels of CDR ≥ 0.5 are not included here.
|  |  | analysis |  |  |
| --- | --- | --- | --- | --- |
| participants<br>(female) | 696<br>(419, 60.2%) | 41<br>(19, 46.3%) | 174<br>(85, 48.9%) | 112<br>(56, 50.0%) |
| scans<br>(female) | 1'927<br>(1'188, 61.7%) | 167<br>(84, 50.3%) | 309<br>(151, 48.9%) | 180<br>(91, 50.6%) |
| age at scan<br>(years, mean<br>+-SD and range) | 69.0 +/- 9.3,<br>(42.7- 97.0) | 74.0 +/- 8.2<br>(54.0-94.4) | 76.1 +/- 7.2<br>(51.7-94.4) | 74.2 +/- 8.4<br>(50.3-95.6) |

### SBA metric computation and normalization

All MRI scans were processed with Ubuntu Linux 22.04.3 LTS on a Dell Precision 7920 workstation with the following specifications. CPU: two Intel Xeon Gold 6148, each one equipped with 20 cores and 2.4 GHz processor base frequency, RAM: 256 GB, GPU: one NVIDIA GeForce GTX 1080 with 8 GB memory. SBA metrics were derived from FreeSurfer (Dale et al., 1999; Fischl et al., 1999a, 1999b; Fischl & Dale, 2000), version 6.0.0 and DL+DiReCT (Rebsamen et al., 2020, 2023a) using default parameters. Results were exported in tabular form using the Desikan-Killiany atlas (Desikan et al., 2006). Because the current implementation of DL+DiReCT does not provide other SBA metrics, only the cortical GMV, mean and standard deviation of the CTh were included in our study. Structures with bilateral representations were used to compute an asymmetry index. In summary, for both processing pipelines, each scan yielded a total of 358 ‘raw’ measurements:

- subcortical volumes: 8 structures (thalamus proper, caudate, putamen, pallidum, accumbens area, hippocampus, amygdala and ventral diencephalon) on 2 hemispheres plus 8 asymmetry indices
- 3 volumes of midline structures (brain stem, 3rd and 4th ventricles)
- cortical regions of the Desikan-Killiany atlas: 3 metrics for 34 regions on 2 hemispheres plus 3×34 asymmetry indices
- brain lobes: volumes for 6 lobes (frontal, parietal, occipital, temporal, cingulate and insula) on 2 hemispheres plus 6 asymmetry indices. Mean and standard deviation of CTh were not included here, since a size-weighted lobar aggregation requires an estimate of the CSA, which is currently not provided by DL+DiReCT.
- brain hemispheres: left/right cortex volume and mean CTh plus asymmetry indices
- whole brain: estimate for intracranial volume (ICV).

In addition to the ‘raw’ metrics, we used ‘normalized’ variants to account for the fact that most metrics vary with brain size. All volumes were scaled to the mean ICV of the normative dataset. Mean and standard deviation of the CTh were instead scaled isometrically according to ICV^(1/3) to respect the geometry of the cortex as a thin two-dimensional sheet, which is folded into three-dimensional space, see (Rummel et al., 2017, 2018) for details. As estimates for ICV we used the Estimated Total Intracranial Volume (eTIV) for FreeSurfer and an exhaustive volume sum of all intracranial segmentations for DL+DiReCT. Since ICV normalized by itself has the same value for all scans and asymmetry indices do not change under the normalization procedure, we obtained 239 additional ‘normalized’ metrics.

### Uniform age sampling

Deviating from the original procedures described in detail in (Rummel et al., 2017, 2018), each one of the 597 SBA metrics (raw plus normalized) extracted from all 1’927 scans with CDR=0 was resampled 100 times by creating 10-bin-histograms of the participant age and drawing n_min_ random samples from each bin, where n_min_ was the smallest bin count. For uniform age distributions, this procedure has no effect, whereas non-uniform age distributions are rendered approximately uniform.

### Normative modeling

Normative models were built for each software and metric independently according to the pipeline of (Rummel et al., 2017, 2018). In brief, low order polynomials were fitted to the 100 resamples of the SBA metrics of our HCs as a function of age. The degrees of the fit polynomials were adapted for each of the resamples separately by increasing from zero until the reduction of residual variance became insignificant (nested F tests). To exclude overfitting, the maximum degree was set as the odd number 2*floor(ln(n/10)+1)-1, where n is the available number of samples (Rummel et al., 2010). For example, when using all 1’927 scans with CDR=0, the maximal allowed degree was 11. The polynomial age trend and prediction intervals were finally computed from the average of all fits to the 100 resamples. Before each of these fits, outliers were removed based on whether they exceeded the 25^th^ or 75^th^ percentile of the distribution by more than 1.5 inter quartile ranges. This procedure was repeated for the fit residues, before a final age fit to the retained data points was generated in the same manner.

### Evaluating patient data against the normative models

With the normative age models available, we applied them to patient scans and compared their fit residues to the distribution in the HCs. Covariates other than age (i.e. sex, scanner and scanning protocol) were accounted for by selecting matched subgroups before computing statistics. Since this matching yielded variable group sizes, the probability P of finding a fit residue of the observed size was calculated accounting for the distribution in the matching HCs and the uncertainty of the measurement, see Rummel et al. (2017, 2018) for details. To account for metric and region specific measurement uncertainties, these were estimated based on repeated scans of the same HC within an age change of less than 10%. Note that compared to same-session rescans under identical conditions, this estimate yields only an upper bound of the true uncertainty. Finally, log10(P), signed positive/negative for larger/smaller than expected fit residues, were used as the central objects to decide whether a regional metric was classified as statistically normal or abnormal. To detect a statistical anomaly, a significance threshold was set to q=0.01, equivalent to -log10(P) > 2.

### ROC curves

To explore the separation of the AD and HC groups, receiver operator characteristics (ROC) and areas under the curve (AUC) were estimated separately for DL+DiReCT and FreeSurfer. The percentage of abnormal metrics per scan (p-values below 0.01) was taken to assess scans as a whole. To focus on brain regions that are known to be affected in AD patients, a similar analysis was repeated for the signed log10(p) values of the CTh of the entorhinal cortex and the hippocampal GMV.

### Comparing spatial patterns

Significance maps and anomaly maps of individuals or groups were compared using normalized L2-distances. L2 was used instead of the Pearson correlation coefficient, because the latter is invariant to shift and scale, which we want to account for when ranking individual maps relative to a template.

### Evaluating and cleaning the normative dataset

The normative dataset was evaluated with a subject-wise leave-one-out cross-validation (LOOCV) study, building normative polynomial models under exclusion of a specific HC (all sessions and repeated scans) and testing all scans of the excluded subject against that model, similar to what was described above for patients. To test for normality of fit residues in our LOOCV, Shapiro-Wilk tests were performed on each of the 597 metrics separately. To test whether the number of detections during the LOOCV was abnormally high over all subjects and ‘raw’ metrics, we performed a binomial test with the number of positives given by the number of anomaly detections, the number of samples given by the number of metrics times the number of scans and the expected fraction of random outliers given by the significance threshold q=0.01.

To clean our normative models from scans with artifacts or potential pathologies before final application, the LOOCV analysis was in addition used to identify anomalous scans separately for the DL+DiReCT and FreeSurfer pipelines and remove them from the normative datasets. We considered scans as not (entirely) normal if they yielded p-values lower than q=0.01 for 18 or more out of the 358 raw metrics (5% of metrics). As a final step, the LOOCV procedure was repeated after cleaning of the normative dataset. The patient evaluation described in the previous paragraph was done against the clean normative dataset.

## Results

We first present results from the AD patient evaluation, followed by some more technical results regarding LOOCV evaluation of HC subjects and dataset cleaning required before patient evaluation.

### Application to AD patients

The AD dataset consisted of 209 scans with CDR >= 1.0. When using the clean HC dataset to evaluate AD scans, both processing pipelines indicated increased proportions of anomalous scans (i.e. scans with more than 18 abnormal raw regional metrics out of 358, equivalent to 5%) in the AD dataset compared to HC. DL+DiReCT resulted in 117 anomalous AD scans (56.0% of all AD scans, compared to 1% in the clean HC dataset), whereas more scans were classified as anomalous using Freesurfer (129 AD scans, 61.7%, compared to 0.8% in the clean HC dataset). Details regarding anomaly detection rates in HC can be found in the section “Anomaly detection in healthy controls (LOOCV)” below.

Figure 1 compares the regional percentage of statistical CTh anomalies detected by ScanOMetrics in individual scans (with significance P<q=0.01, not corrected for multiple comparisons) in patients with AD as well as in the cleaned HC dataset. The patterns of preferred anomaly detection are remarkably similar between both processing tools and symmetric with respect to hemispheres. Comparison of the CTh reduction map in patients with AD (third row) with the effect size map of a direct statistical comparison between the AD and HC groups (Cohen’s d, bottom row) displays remarkable agreement of the temporo-parietal atrophy patterns. In patients with AD, reduction of regional mean CTh is detectable in up to 28% of individual scans with a strong regional preference for the bilateral entorhinal and fusiform cortex as well as in the precuneus and supramarginal gyrus. In the frontal lobe the CTh reduction is weakest. For HCs the peak percentage of detected CTh reductions is only in the order of ∼1.3%, i.e. twenty-fold reduced when compared to patients with AD. Increase of CTh is also observed in up to ∼4.5% of patients with AD, with peak in the bilateral medial orbito-frontal gyrus and cuneus.

**Figure 1:**
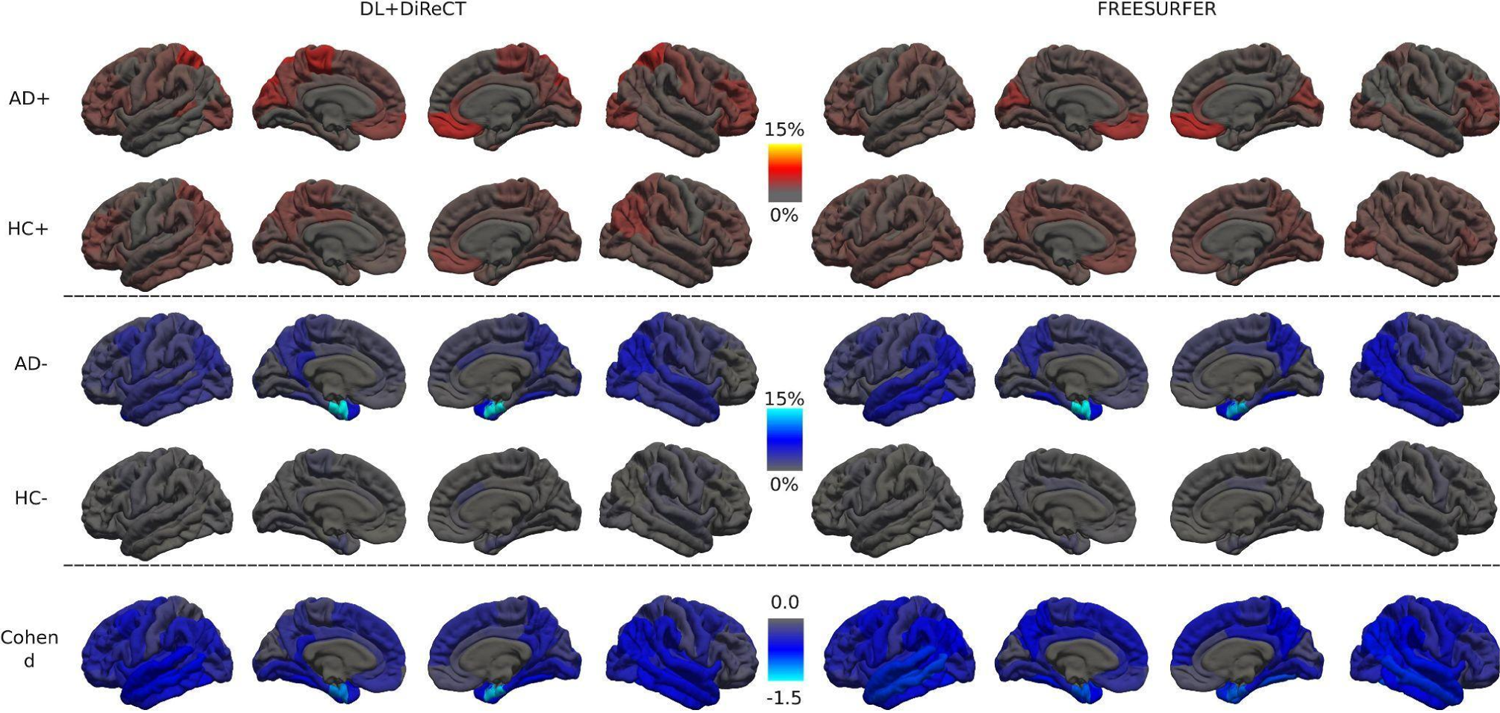
Percentage of CTh anomalies in the AD and HC groups, detected with ScanOMetrics using both processing tools. AD patients with established dementia (CDR>=1, 209 scans) are shown in rows 1 and 3, results of the LOOCV in cleaned non-convertering HCs (CDR=0, 1’828 scans) in rows 2 and 4. Deviations towards larger (rows 1 and 2, red-to-yellow colormap) and smaller (rows 3 and 4, blue-to-white colormap) than expected CTh are collected separately. The bottom row shows the effect size (Cohen’s d) when contrasting the entire AD and HC groups. Positive effect sizes did not occur in this comparison.

When stratifying scans from patients with AD by the clinical dementia rating (CDR), a clear progression pattern with spread of atrophy along the temporal, parietal and eventually frontal lobe regions is revealed by the CTh anomaly maps, see Figure 2. Using DL+DiReCT, abnormal mean entorhinal CTh is detected already in about 24.8% of patients with CDR=0.5 (N=416), which progresses to 47.2% of patients with CDR=1 (N=159), and 46.0% of patients with CDR>=2 (N=50). In contrast, using FreeSurfer, abnormal thickness is detected in 17.6% of CDR=0.5 patients, 30.2% of CDR=1 patients, and 32.0% of CDR>=2 patients. For CDR>=1, CTh reduction becomes visible in the precuneus and supramarginal gyrus as well. For cases with CDR>=2 also the fusiform gyrus (20% of cases for DL+DiReCT and 34% of cases for FreeSurfer) and the lateral temporal lobes become affected. In the OASIS3 dataset an increase of CTh in the bilateral medial orbito-frontal gyrus is observable and associated with increasing CDR, an effect which is clearer visible with FreeSurfer than with DL+DiReCT.

**Figure 2:**
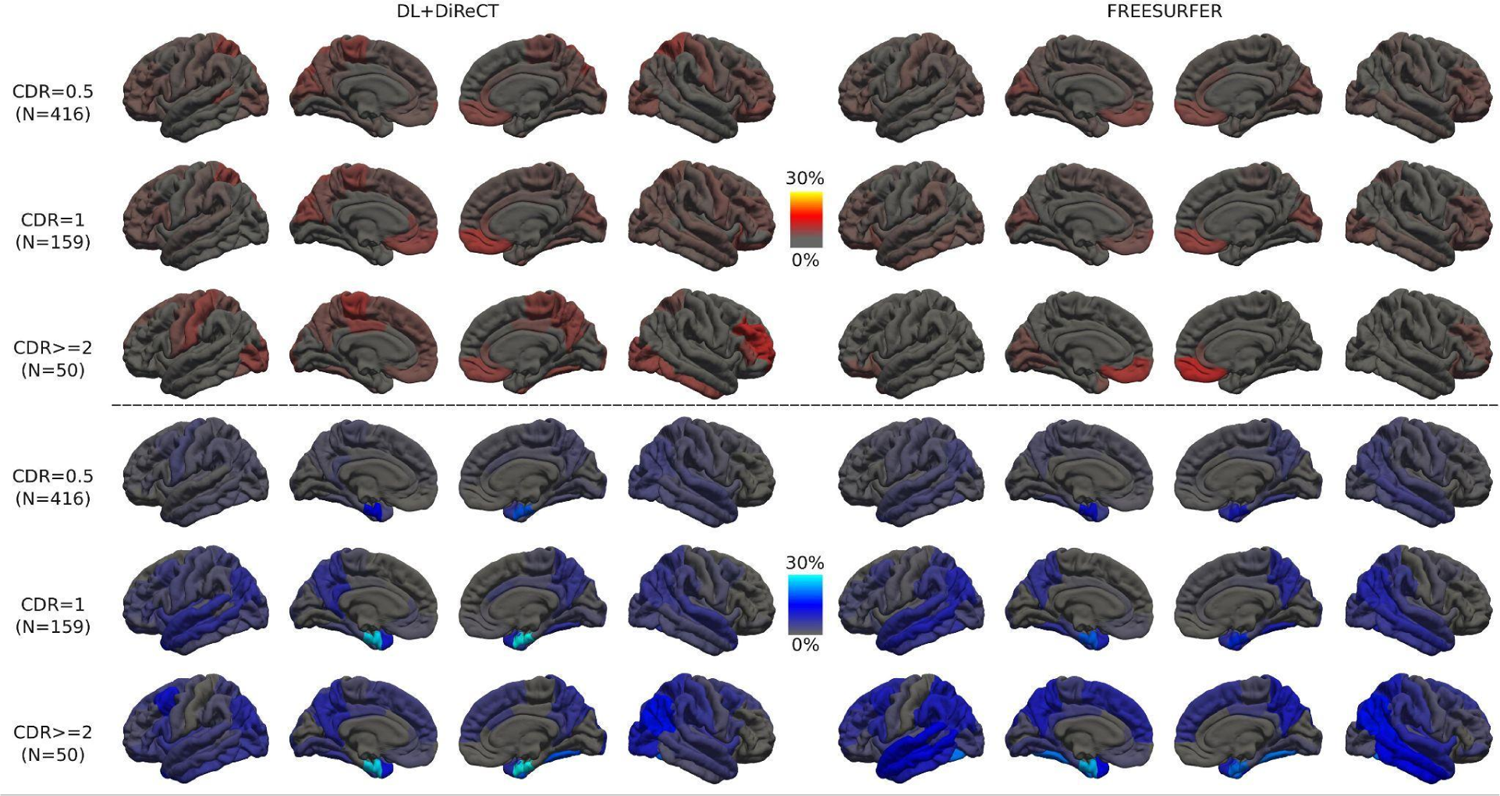
Percentage of CTh anomalies detected by ScanOMetrics in patients with AD, stratified by cognitive impairment levels at scan time (CDR=0.5: rows 1 and 4, CDR=1: rows 2 and 5, and CDR>=2: rows 3 and 6). The upper half depicts CTh increase, while the lower half shows progression of CTh reduction. Mind that the color scales are different from the ones used in Figure 1.

Figure 3a compares normalized L2-distances between ScanOMetrics’ individual significance maps of all scans with CDR>=0.5 and the average significance map of all scans with CDR>=2, which was used as a template for clear AD. Estimates from DL+DiReCT and FreeSurfer were found highly correlated (r=0.87, p<1e-16). Figure 3b shows histograms of the L2-distances, separately for DL+DiReCT and FreeSurfer, grouped by increasing CDR and revealing a negative association for both tools. Figure 3c presents examples of individual significance maps for five different scans. Selection was made based on quantiles of the normalized L2-distances shown in panels a and b. Interestingly, the scan closest to the CDR>=2 template was the same one for DL+DiReCT and FreeSurfer (corresponding to the lowest left datapoint in Figure 3a, a scan with CDR=0.5). Figure 3c illustrates at the same time how diverse significance maps can look like in different patients (top and middle row), how similar anomaly detection can be for both software tools (bottom row), and how loosely individual clinical scores and corresponding significance maps in structural MRI scans can be related (the scan closest to the AD template generated from CDR>=2 cases has a CDR of only 0.5).

**Figure 3:**
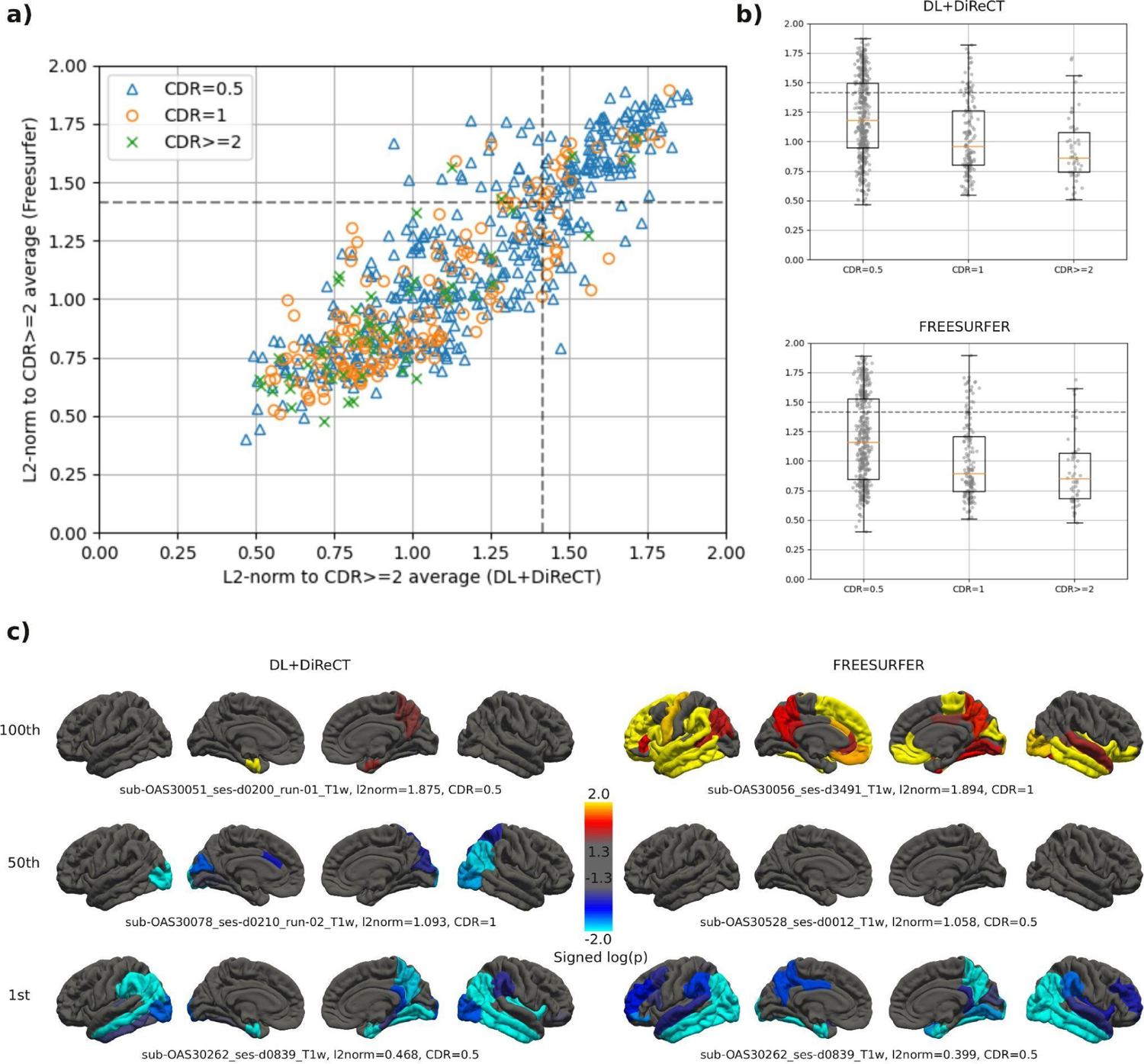
Normalized L2-distances between individual significance maps and a template (i.e. the average map of all scans with CDR>=2). Significance maps are log10(p) maps with negative sign for CTh reduction and positive sign for CTh increase. Normalized L2-distances range from 0 for identical maps to 2 for antisymmetric maps, with sqrt(2) indicating orthogonal maps (marked by dotted lines in panels a and b). a) Correlation between DL+DiReCT and FreeSurfer, individual CDR scores are symbol/color coded. b) Grouping by CDR separately for both software tools. c) Significance maps in individual scans, selected according to their L2-distance. Scans on the 1st row are the 100th percentiles in the distributions (i.e. highest distance to the reference), while the lowest row are the most similar to the group average. In the lowest row the same scan was selected for both DL+DiReCT and FreeSurfer, and corresponds to the data point closest to the origin in panel a).

In agreement with published results (van Hoesen et al., 1991; Gómez-Isla et al., 1996; Juottonen et al., 1999; Du et al., 2001; Price et al., 2001; Mueller et al., 2010; Devanand et al., 2012; Igarashi, 2023), the bilateral entorhinal gyrus was identified as one of the earliest visible and most prominent deviations in patients with AD from the normative model, see Figures 1 and 2. Hippocampal volume has also been reported to be prominently atrophic in AD (Juottonen et al., 1999; Du et al., 2001; Sluimer et al., 2008; Devanand et al., 2012). In Figure 4 we display the mean normalized volume of the hippocampus (as provided as ScanOMetrics output based on DL+DiReCT estimates) for the scan with the highest individual similarity with the AD group (i.e. the lowest row in Figure 3c) and compare with the point cloud of the cleaned normative dataset. Hippocampal volume is in the order of only 2.5 ml on both hemispheres, much below the 95% prediction interval [3.2 ml, 4.5 ml] estimated from our HCs at the same age. Furthermore, the hippocampal volume was found to decrease bilaterally from the first to the second scan available for this patient. Atrophy rate for the average volume of both hemispheres was 12.4% over a period of 2.2 years. When using Freesurfer (see Supplementary Figure S1), the estimated atrophy rate was lower (6.7% over 2.2 years).

**Figure 4:**
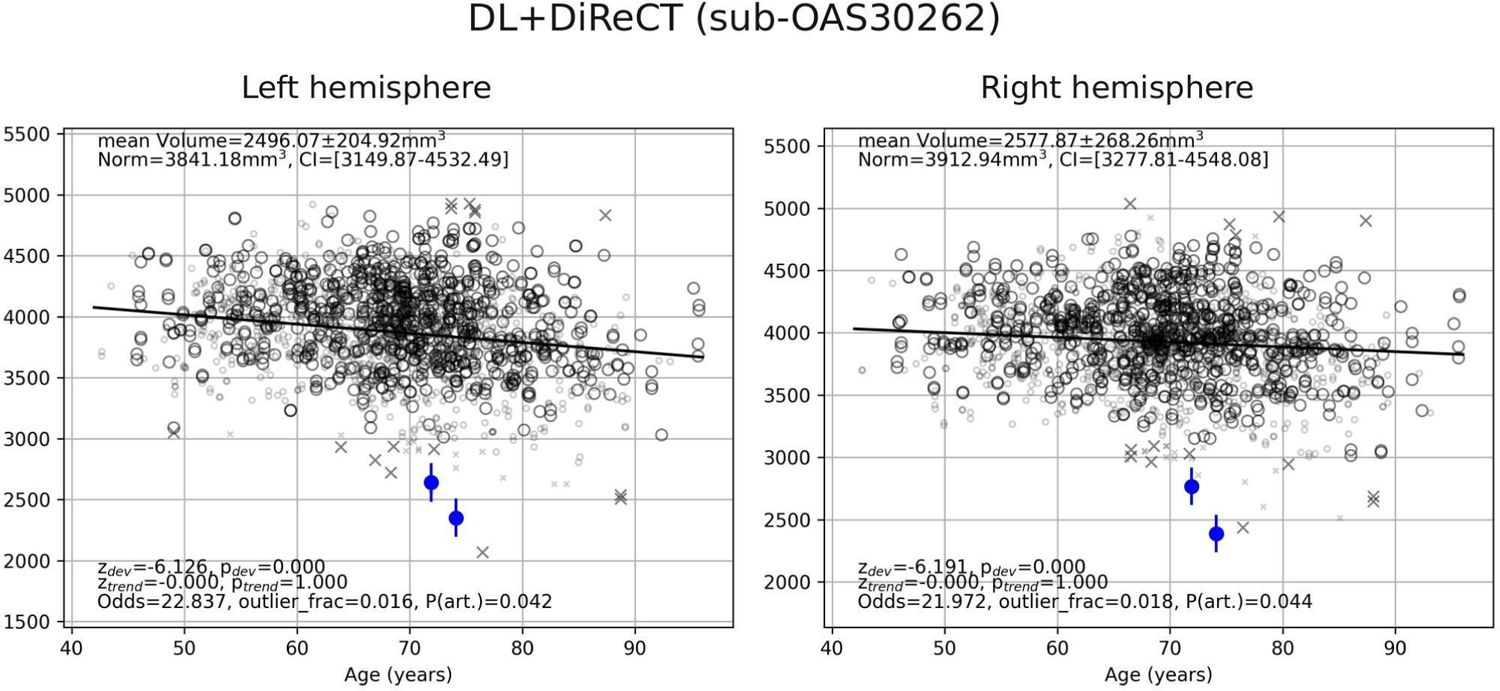
Age dependence of the brain size normalized volume of the hippocampus, as displayed by ScanOMetrics (volume estimates by DL+DiReCT). The corresponding data derived from FreeSurfer is available in our Supplementary Figure S1. Similar results were observed for the normalized CTh of the entorhinal cortex (not shown).Patient data (blue) are the two scans of the participant closest (i.e. had the smallest L2 norm) to the AD group’s average significance map (log10(p) maps for the second scan are shown at the lower left section of Figure 3c). Symbols in black represent the HCs used to build the cleaned normative dataset. Crosses are estimates flagged during outlier removal and did not contribute to statistics. Large symbols match the patient scans regarding sex, MRI scanner type and scanning protocol, whereas small symbols differ in at least one of these characteristics. Fully drawn lines indicate the fitted age trajectory of the normative models. Significance of statistical comparisons and the reliability of the measurement (see Rummel et al., 2017, 2018 for details) are reported in the lower left corners of the panels. Values reported in the upper left corner are the subject average across time points, along with the expected value from normative data and its prediction interval.

We used the 87 MRI scans of the 41 subjects that were excluded from building the normative models (conversion from CDR=0 to CDR>=0.5) to investigate the change of thickness over time in more detail. To focus on the clinically relevant question of early atrophy detection, we restricted this analysis to participants where a scan with CDR=0 was available, excluding any progression between higher CDR levels. Difference maps of mean regional CTh (ICV normalized, later scans minus baseline always, regardless the associated CDR values) were averaged over all scan pairs of the selected 41 subjects and are displayed in Figure 5. Similar to Figure 2, where progression is displayed by grouping according to CDR, the most prominent atrophy progression over time occurred in temporo-basal brain regions, like the entorhinal, parahippocampal, fusiform and inferior temporal gyrus, where mean CTh reduced up to 0.1 mm, a change equivalent to the expected reduction of whole brain mean CTh in 25 years of healthy aging (Lemaitre et al., 2012). Also remarkable is the relative sparing of the somato-sensory cortex from atrophy progression (Thompson et al., 2003; Lerch et al., 2005; Fennema-Notestine et al., 2009; Frisoni et al., 2010; Rebsamen et al., 2020), which becomes most transparent in the left precentral gyrus in Figure 5 but can be identified in individual scans of Figure 3 and in the percentage maps of Figures 1 and 2 as well.

**Figure 5:**
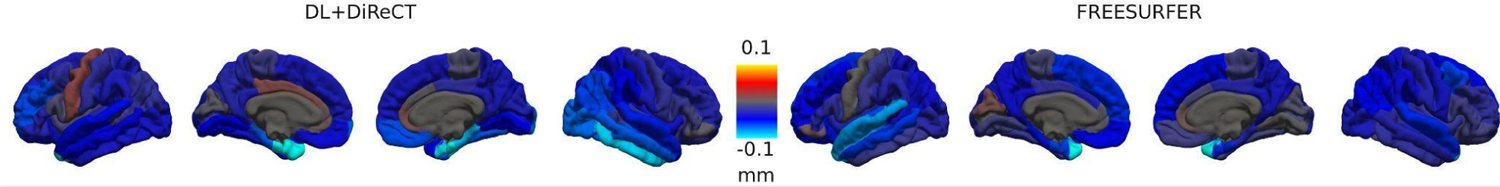
Average change in mean normalized regional CTh in subjects converting between CDR=0 and CDR>=0.5. In contrast to Figures 1, 2 and 3c changes are measured in millimeters here.

### Scan classification

Classifying scans as AD/abnormal based on the percentage of metrics with p-value below 0.01 lead to AUCs of 0.76 for DL+DiReCT and 0.72 for FreeSurfer (Figure 6). Sensitivity and specificity were the closest to the top-left corner when using a threshold of 1.04% for DL+DiReCT (FPR=0.29, TPR=0.69) and 0.62% for FreeSurfer (FPR=0.35, TPR=0.69).

**Figure 6:**
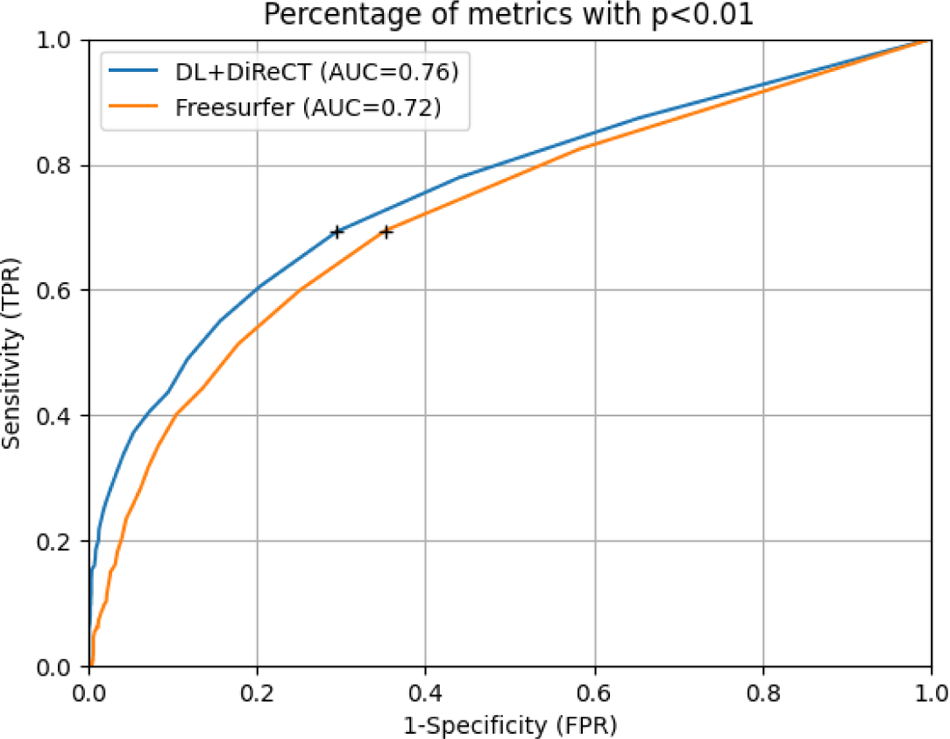
Receiver operator characteristics (ROC) for classification of scans into AD and HC, based on their percentage of abnormal metrics. Patients with AD were evaluated against the normative model of the clean HC dataset and all HC scans against this subject’s LOOCV model. Black crosses show thresholds for which the points on the ROC (sensitivity and 1-specificity) were closest to the top-left corner.

Instead, using a fixed threshold of 5% abnormal metrics to label a scan as abnormal (i.e. the threshold used to clean the original dataset) lead to FPR=0.03 and TPR=0.28 for DL+DiReCT, while the rates were 0.04 and 0.20 for Freesurfer. Similar results were obtained for the attempt to classify scans based on the signed log10(p) value of the CTh of the entorhinal cortex (DL+DiReCT slightly better, see Supplementary Figure S5) or of the hippocampal GMV (FreeSurfer slightly better). Both tools had the same discriminant power when using the suitable metric (AUC=0.75).

### Within-subject reproducibility/homogeneity

Supplementary Figure S2 shows an example of CTh deviations in the patient with CDR>=2, who had the largest number of scans (OAS30902, four rescans during the same session). The figure consistently shows atrophy patterns in the right parietal and temporal lobe, as well as the characteristic reduction in CTh in the entorhinal cortex, extended to the lingual gyrus. Interestingly, both FreeSurfer and DL+DiReCT indicate increased CTh in several regions in the first two rescans. Visual inspection of these scans showed reduced image contrast, presumably due to patient motion, explaining the need to acquire two additional scans, which had better image quality.

Reproducibility of CTh patterns across the whole OASIS3 dataset was assessed. Subject-wise distances of the CTh significance maps between rescans of the same participant were estimated by calculating the normalized L2-distance between signed log10(p) maps of mean CTh estimates (brain size normalized). HC maps were taken from the LOOCV analysis, whereas AD maps were taken from their evaluation against the clean normative dataset. When using DL+DiReCT, the distance between significance maps of repeated scans was lower in patients with AD (L2=0.39 +/- 0.26, median +/- standard deviation) than in HCs (L2=0.42 +/- 0.19, p=0.05 in a Wilcoxon rank sum test to account for the large skewness of both distributions). For Freesurfer, there was no significant difference between AD and HC (AD: 0.56 +/- 0.26; HC: 0.56 +/- 0.20; p=0.66). Repeated significance maps were significantly closer for DL+DiReCT than for FreeSurfer (p=9.2e-57 in Wilcoxon signed rank test on AD maps, and p=2e-309 on HC maps).

### Processing times

On our hardware the processing time for one MRI scan was 9h20m +/- 2h50m (mean +/- standard deviation) with FreeSurfer (running on CPU only), and 23m59s +/- 4m30s with DL+DiReCT. This value was split into 1m55s +/- 13s for segmentation on the GPU and 22m04s +/- 4m25s for CTh estimation with DiReCT (Das et al., 2009; Avants et a., 2014) on the CPU. Fitting the clean normative models on all subjects took 8m17s for FreeSurfer and 7m04s for DL+DiReCT. Time required for evaluation of a single scan against a normative model was 1.91 +/- 0.57 seconds for FreeSurfer and 1.93 +/- 0.58 seconds for DL+DiReCT.

### Cleaning the normative models

Among the 1’927 HC scans, that were initially used for normative modeling, 99 scans were flagged as anomalous in the LOOCV analysis (i.e. more than 18 metrics with P<q=0.01), using either DL+DiReCT or FreeSurfer. Figure 7 shows the cumulative distributions (left) and probability densities (right) of the fraction of abnormal metrics per scan, and details the number of rejections for both pipelines. Regions that contributed to the large number of anomalies in these 99 scans were widely distributed over the entire cortex, see upper part of Supplementary Figure S3. Furthermore, visual inspection of the scans with the largest number of deviant metrics (see Supplementary Figure S4) revealed the following potential causes for outlier detection:

low image quality, mainly due to susceptibility artifacts in the mouth region (rows 1 and 2); likely caused by dental implants (Chockattu et al., 2018)
prominent lateral ventricles and/or enlarged CSF space suggestive of atrophy (e.g. subject OAS30662, row 3; about 20% of DL+DiReCT’s metrics were flagged as abnormal with respect to the expected values at the age of the subject, despite a reported CDR score of zero)
blurring and ringing artifacts due to patient motion during the scan (rows 4 to 6)

**Figure 7:**
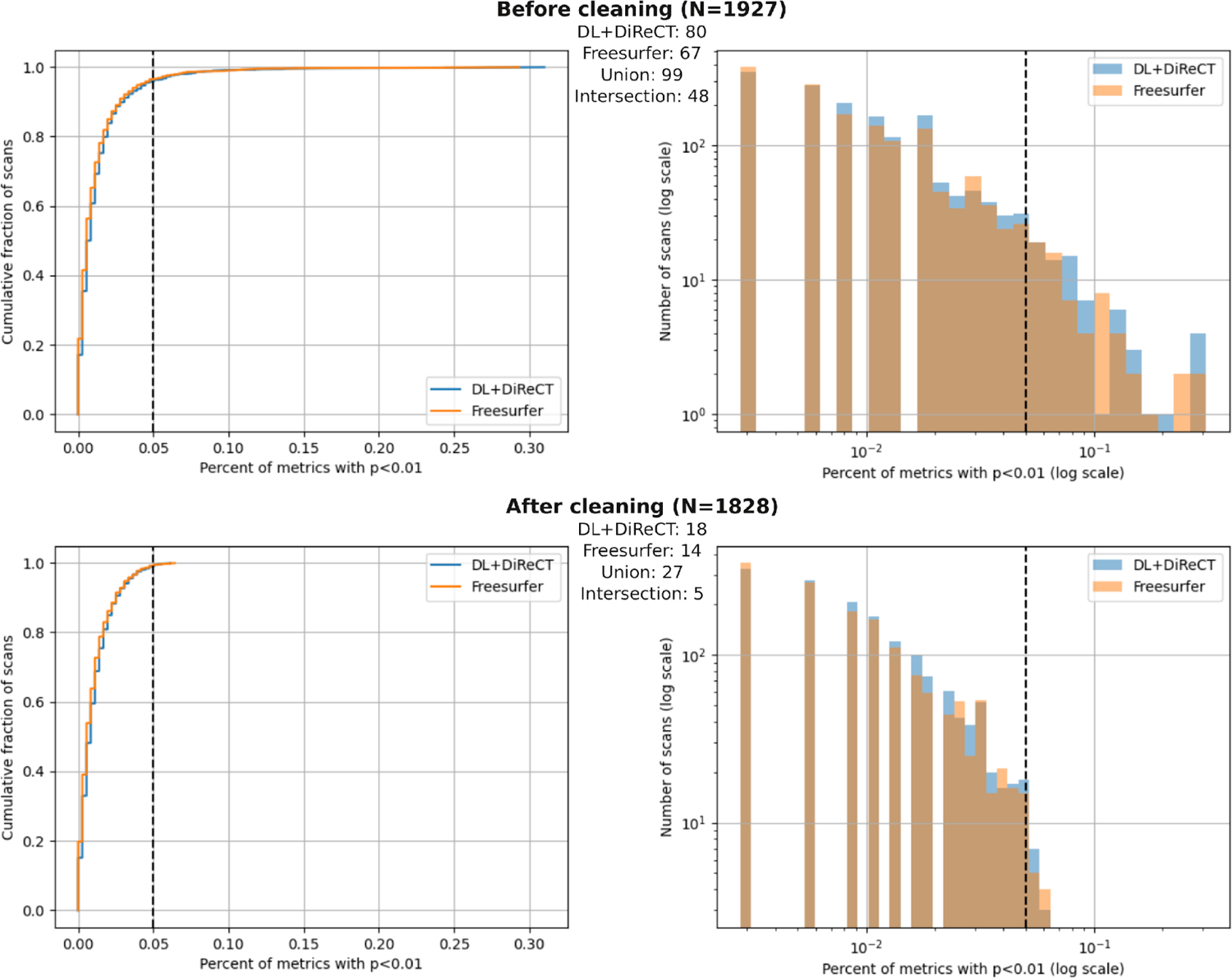
Distribution of the abnormal fraction of metrics per scan, before (top) and after (bottom) cleaning the normative dataset. Numbers in bold correspond to the total number of scans before and after cleaning. Numbers in smaller font correspond to anomalous scans in the dataset, as detected when using either DL+DiReCT or FreeSurfer. Cleaning consisted in removing the 99 scans detected as anomalous by either one of the two software tools.

The 99 HC scans with large number of anomalies (5.15%) were removed before our final normative models were built from the of 1’828 remaining HC scans. These “clean HC datasets” were subjected to a final LOOCV and used for all subsequent analyses of scans from patients with AD. In these models the number of HC scans flagged as anomalous by either DL+DiReCT or FreeSurfer decreased to 27 (1.48% of HC scans, p_binom_=0.045 for fraction 1%). Especially for DL+DiReCT the anomalies in these scans were much more regionally specific than before (lower part of Supplementary Figure S3). Shapiro-Wilk tests indicated that almost all metrics had non-normally distributed fit residues, with only minor improvements due to the cleaning procedures (decrease from 91% to 85% for DL+DiReCT and from 90% to 84% for FreeSurfer regarding the 358 raw metrics, and from 93% to 90% for DL+DiReCT and 98% without change for FreeSurfer regarding the 240 brain size normalized metrics).

### Anomaly detection in healthy controls (LOOCV before cleaning)

We assessed specificity of our approach to regional anomaly detection by running a subject-specific leave-one-out cross-validation (LOOCV) on the whole HC dataset. When considering the full set of tests made (358 raw metrics times 1’927 scans, yielding 689’866 p-values), and using a significance threshold of q=0.01, processing the HC dataset with DL+DiReCT resulted in 8’911 significant p-values (1.30%, which is slightly but significantly higher than the expected 1%, p_binom_<1e-16). When considering an alternative q=0.05, only 4.44% of p-values were significant, which was lower than the expected 5%. Processing data with FreeSurfer, using q=0.01 or q=0.05 resulted in 1.16% and 4.13% of significant p-values, respectively, which more or less resembled the numbers reported by Rummel et al. (2018) for a completely different dataset. Similar to the suspicion raised there, the reason for this observation might be due to the residues of the polynomial age fits not being normally distributed (Shapiro-Wilk tests) for the vast majority of metrics in our LOOCV, regardless of whether using DL+DiReCT (305 metrics out of 358 were not normally distributed) or FreeSurfer (299 of 358 non-normally distributed sets of residuals), and independently of using uniform subsampling or not.

### Features of the cleaned normative models

After cleaning, DL+DiReCT yielded 18 anomalous scans with more than 5% of regions detected as anomalies (0.98% of the 1’828 scans), while FreeSurfer detected 14 scans (0.77% of scans). Union of both sets yielded 27 scans (1.48%), and the inter section 5 (0.27%). Values are reported in the lower part of Figure 7, and correspond to scans on the right of the dotted lines in the cumulative distribution and histogram.

Since mean age was different between our participants with CDR=0 and CDR>=1 (see Table 1, p=3.3e-13 in a t-test), fitting age models and working with residues rather than with the original metrics was appropriate. Normative models for metrics estimated with DL+DiReCT had degree d=0 (constant) in 55% of the fits, d=1 (linear) in 42% of the fits, and d=2 (quadratic) in 3% of the fits. No higher degree was selected. When using FreeSurfer, the degree was d=0 in 25.7% of the fits, d=1 in 70.3% of the fits, d=2 in 3.9% of the fits, and d=3 (cubic) in 0.1% of the fits.

We performed 240 one-tailed F-tests (once for either direction) for different residual variance when fitting normative models to brain size normalized metrics calculated with DL+DiReCT or FreeSurfer. Testing smaller variance for DL+DiReCT than for FreeSurfer and performing FDR correction to account for the many comparisons, 56 of the 240 metrics were significant on level q<0.01. Most of these were either mean normalized CTh (16 of 34 on the left hemisphere and 15 on the right) or its standard deviation (8 on the left and 11 on the right). In contrast, only seven residual variances of normalized GMV were smaller for DL+DiReCT than for FreeSurfer, among which five were subcortical regions. Testing in the other direction, 97 metrics showed smaller residual variance for FreeSurfer than for DL+DiReCT. Among these, 78 were cortical and subcortical volumes, whereas only 2 (4) were mean and 6 (7) were standard deviations of normalized CTh on the left (right) hemisphere.

## Summary and Discussion

In this paper we have compared identical metrics derived from two brain morphometry software tools, i.e. DL+DiReCT (Rebsamen et al., 2020, 2023) and FreeSurfer (Dale et al., 1999; Fischl et al., 1999a, 1999b; Fischl & Dale, 2000), regarding their use in the context of ScanOMetrics, an open-source pipeline for normative modeling and detection of statistical anomalies (Rummel et al., 2017, 2018). ScanOMetrics processing is supposed to detect abnormal regions in individual MRI scans, which may support neuroradiological assessment of the cases with respect to many clinical questions. An implementation of ScanOMetrics in Python3 has been made publicly available to the community as open source software. Together with the public availability of the used OASIS3 dataset, ScanOMetrics tutorials available online and the normative models used in the present work (specific for OASIS3), this makes our results completely reproducible.

Our main findings are that regardless of the software used for extraction of the metrics, in patients with Alzheimer’s disease (AD) anomaly detections were up to twenty-fold more frequent than in healthy controls (HC). Cortical thickness (CTh) anomalies were mainly detected in regions that are known as predilection areas of cortical atrophy in AD and progression of the atrophy pattern with clinical dementia rating (CDR) was clearly observable with both methods. DL+DiReCT provided CTh results more than 15 times faster than FreeSurfer.

### Origin of statistical brain anomalies

Detected statistical anomalies may have at least three origins, which influence their differential statistical properties. First, regional metrics of brain shape can artifactually be detected as abnormal. These detections depend on the measurement uncertainty of the metric and the image quality of the scans. They have spatial predilection regions, which can be identified by studying scan-rescan variabilities (see e.g. Rummel et al., 2018; Rebsamen et al., 2020). In ScanOMetrics the artifact probability and odds for valid vs. artifactual detections are reported (see text elements in Figure 5) to guide the user’s judgment of reliability. Second, different individuals have different brain shapes, which yields highly reproducible deviations from the expectation, see Section “Within-subject reproducibility/homogeneity” and Rummel et al. (2017, 2018). Where these deviations are strong enough, they can trigger subject specific anomaly detections. Finally, brain pathologies yield anomaly detections as well, which are often concentrated in brain regions that have been revealed as disease specific alterations of brain shape in large morphometric group studies in the past.

Importantly, when investigating an individual MRI scan (as is often the case for diagnostic purposes), all three sources contribute to detections of statistical anomalies, but only the last category is relevant to answer clinical questions. In consequence, the pattern of detections (e.g. spatial extent of alterations of brain structure, like e.g. atrophy) in an individual almost never matches disease specific patterns as described in the literature or derived by group assessment exactly. While often centered in these predilection areas, detections depend on image quality and usually reach beyond these regions, see Figure 3c for an illustration of the variability in patients with AD of the OASIS3 dataset. However, when pooling the detections made in many individuals over groups representing the same clinical condition like in our Figures 1 and 2, the first two causes for anomalies have a chance to level out and the expected group patterns usually become visible more clearly. Similarly, when pooling several scans of the same subject, primarily the subject-individual anomalies would become more clearly visible, whereas the same is true for disease-specific patterns only if they remain stable over the observation period.

### Atrophy patterns in individual patients with Alzheimer’s disease

Our hypothesis (4) was that ScanOMetrics yields a much higher rate of detected anomalies in patients with AD than in HCs when using a leave-one-out cross-validation (LOOCV). Indeed, independently from the used software, about 60% of scans were rated as abnormal in the AD group, compared with only ∼1% in the cleaned HC dataset. Comparison of rows 3 and 4 in Figure 1 shows the associated rates of atrophy detection in individuals. Particularly in the bilateral entorhinal gyrus (see e.g. Gómez-Isla et al. 1996, or Mueller et al., 2010), but also in parieto-temporal brain regions, the rate of detected significant reductions in CTh was elevated in patients with AD up to twenty-fold. Regarding detected regional increase in CTh, the difference between AD and HC scans was much less pronounced (rows 1 and 2).

The spatial pattern of detected CTh reductions was consistent with the temporo-parietal predilection areas of atrophy in patients with AD obtained from a group comparison in the same data (see Figure 1, rows 3 and 5) or existing literature like (Whitwell et al. 2009, Harper et al. 2017, Ferreira et al. 2017), confirming the second part of our hypothesis (5). A similar correspondence between pooled detections in individuals and results of a group study was recently found by Verdi et al. (2023).

Figure 2 reveals that the rate of detected brain atrophy (rows 4 to 6) increased with clinical dementia rating (CDR). Using DL+DiReCT, the mean CTh of the bilateral entorhinal gyrus was detected as significantly reduced already in one fourth of patients with CDR=0.5, a value that almost reached half of the scans in cases with CDR>=1. Using FreeSurfer the progression with CDR was observable as well, but detection rates were lower (i.e. only about one sixth for CDR=0.5 and in one third in CDR>=1). A similar association between atrophy detection in scans of patients with AD and their total scores from the Mini-Mental State Examination (MMSE) has been observed recently by Verdi et al. (2023). Furthermore, our Figure 2 showcases the posterior-to-anterior atrophy progression reported by Contador et al. (2021).

Regional CTh increase for higher CDR (rows 1 to 3 of Figure 2) was observed only for the medial orbito-frontal gyrus (predominantly on the right hemisphere, more pronounced for FreeSurfer than for DL+DiReCT). Interestingly, this is exactly the region in which increased CTh was detected in individual scans with reduced image quality due to patient motion (see rows 1 and 2 of Supplementary Figure S2 for an example). In addition, Supplementary Figure S3 reveals that the medial orbito-frontal gyrus was one of the more frequently made false positive detections in HCs after cleaning of the normative dataset. Since we do not have any plausible interpretation of the observed CTh increase, we hypothesize this as a plausible effect of an association between CDR and patient motion during the scan.

Compared to the large fraction of entire scans rated as anomalous in patients with AD (∼60% for both software tools), the peak effect of CTh reduction (detectable in the entorhinal gyrus in “only” about half of scans of the general AD group, see Figure 1, row 3) was relatively small, indicating that the anomaly patterns detectable in the individual patient with AD are largely non-overlapping. This observation confirms the first part of our hypothesis (5) and is consistent with recent observations by Verdi et al. (2023), who have also reported widespread detection patterns with only moderate peak proportions of detections in the basal temporal lobes.

### Value for clinical decision support

Normative modeling of healthy brain shape, its development and aging have great potential to support clinical routine assessment of suspected pathologies in neuroradiological MRI exams. It is important to stress that we envision the automated detection of statistical anomalies in individuals (like shown for example in Figures 3 or S2) as a trigger for secondary inspection by the human expert, rather than as an automated disease classification tool. Used as a screening tool for further regional image analysis (Rummel et al., 2017), normative modeling could indeed provide valuable decision support to the neuroradiologist.

In contrast, classifying entire scans as anomalous based on a threshold on the accepted rate of abnormal metrics is not sufficiently reliable. The same is true for classifying scans into AD or HC based on the accepted degree of anomaly in selected regional SBA metrics, which are known as frequently compromised in dementia (like e.g. the hippocampal GMV or the entorhinal CTh). In our study, both approaches lead to areas under the ROC curve below 0.8, without greater difference between the evaluated software tools.

CE-marked and FDA-approved commercial tools for clinical decision support by brain morphometry have meanwhile become available for application in patients with multiple sclerosis and various forms of dementia. Despite formal approval for diagnostic purposes, a deficiency of these tools is that validation, especially in clinical terms, in many cases still is an open topic of research (Pemberton et al., 2021; Mendelson et al., 2023) due to a multitude of factors (Haller et al., 2022; Leming et al., 2023; Hedderich et al., 2023). This is remarkable, since an international survey among practitioners investigating their application of (commercial or scientific) brain morphometry tools has clearly shown that user acceptance is associated with the availability of technical and clinical validation studies (Vernooij et al., 2019).

### DL+DiReCT vs. FreeSurfer

Our comparison between using DL+DiReCT and FreeSurfer for metrics estimation was motivated by the question, whether one of the two methods yielded more stable or more plausible spatial patterns of statistical anomaly detections than the other, see our hypotheses (1), (2) and (3). Our findings show that this question cannot be answered so clearly. In general, the group aggregations in Figures 1, 2 and 5 reveal very similar patterns for both software tools. The lowest row in Figure 3c and Supplementary Figure S2 show that the same can be true for the degree of regional CTh anomaly detected in rescans of an individual patient.

Our hypothesis (1) that fit residues are in general more narrowly distributed for DL+DiReCT than for FreeSurfer could not be confirmed by our study. Rather, this was true only for one fourth of the normalized metrics (56 of 240), whereas 97 showed the opposite behavior. Remarkably, we observed that metrics with smaller residual variance for DL+DiReCT were predominantly thickness measures, whereas volume metrics dominated the group where residual variance was smaller for FreeSurfer. More narrow distribution of CTh fit residues when using DL+DiReCT is in line with recent observations by Rusak et al. (2022), who have found that the DL-based tool is more sensitive and more reproducible at weak synthetic reduction of CTh than FreeSurfer’s cross-sectional or longitudinal pipelines. For GMV the situation is different: DL+DiReCT counts voxels and thus is prone to uncertainties introduced by its voxel-wise hard classification into one of several brain regions or tissue classes. By contrast, FreeSurfer’s GMV estimates are based on the volumes enclosed inside its much smoother surface meshes, which likely explains the more narrow distribution of volume fit residues.

Similarity of deviations from the normative models between rescans of the same participant (assessed by normalized L2-distance of thickness-based signed log10(p) maps) was large in general. This is in line with the observation that ScanOMetrics’ deviations from the expectation are subject specific (Rummel et al., 2017, 2018) and remarkably reproducible, also see Supplementary Figure S2 for an example. Confirming our hypothesis (2), L2-distances of CTh significance maps between rescans were significantly smaller for DL+DiReCT than for FreeSurfer and a similar effect was observed for cortical GMV (not shown). This is consistent with the interpretation that rescan errors and disturbance by artifacts depending on image quality are smaller for DL+DiReCT than for FreeSurfer. The L2-distances were in addition smaller in patients with AD than in HCs when using DL+DiReCT. We interpret this finding as a sign that subject-specific signed log10(p) values derived using DL+DiReCT are small and spatially unspecific for HC subjects, whereas those of patients with AD have additional disease related and spatially specific deviations from the normative model that are larger in size and thus determine the L2-distance.

In Figure 4 we have detected an annual hippocampal atrophy rate of almost 6% in an individual patient using DL+DiReCT, which is in agreement with group estimates found in the literature (Sluimer et al., 2008). Using FreeSurfer the annual atrophy rate was only half as large (see Supplementary Figure S1). This might indicate a higher sensitivity of DL+DiReCT to atrophy progression in the individual, supporting our hypothesis (3). Since sensitivity to atrophy and reproducibility of patterns has mainly been compared for CTh and not for GMV so far (Rebsamen et al., 2020, 2023; Rusak et al., 2022), this hypothesis requires additional investigation in subsequent work.

Processing with DL+DiReCT (<25 minutes) yielded comparable results for mean CTh more than 15 times faster than the full FreeSurfer pipeline. However, DL+DiReCT’s output is drastically reduced, currently focusing on some of the most frequently used SBA metrics of brain morphometry, namely mean and standard deviation of regional CTh, GMV and volumes of some subcortical segmentations. Using the OASIS3 dataset with 1’828 HCs after cleaning, fitting a normative model to the set of regional brain metrics used in this paper took less than 10 minutes. Importantly, this procedure has to be performed only once for each normative dataset. Despite the expectation of statistical post-processing with ScanOMetrics to only depend on the number of metrics and scans and not on the software used for metrics estimation, we observed a minimally smaller processing time for DL+DiReCT than for FreeSurfer. We explain this minor discrepancy by a different number of outliers rejected during the fitting procedures. Application of the normative models to a new case required less than two seconds computation time for both tools, almost three orders of magnitude quicker than the calculation of the metrics with DL+DiReCT and practically not contributing to the entire computation time.

### Outlook

Future work should combine residues from normative modeling with proportional hazard models for AD conversion (e.g. Devanand et al., 2012), AD classifiers (e.g. logistic regression as used in Bobinski et al., 1999, linear discriminant analysis as in Juottonen et al., 1999, support vector classifiers as in Schmitter et al., 2014 and Gupta et al., 2019, random forests or K-nearest neighbor classifiers as in Gupta et al., 2019, probabilistic multi-kernel classifiers as in Popuri, 2020) or disease progression models (Fonteijn et al., 2012; Sivera et al., 2019; Planche et al., 2022; Saint-Jalmes et al., 2023) to thoroughly investigate if improved diagnostic accuracy can be obtained at the subject level. Special care should be devoted to avoid data leakage (Kapoor and Narayanan, 2023), and into addressing the heterogeneity/similarity of atrophy patterns across dementia subtypes.

Atrophy patterns have been shown to differ between early and late onset dementia (Harper et al. 2017), to be similar between AD subjects with and without amnestic clinical syndromes (Whitwell et al. 2009), or even to be undetectable with current methodology in some AD patients (Ferreira et al. 2017). Grouping subtypes with different atrophy patterns might impair the accuracy of clinical decision support models, while splitting datasets in too many groups will reduce statistical power. Further work should explore if individual normative metrics could be of interest for certain dementia subtypes, or if multivariate and disease progression models are required in order to properly classify subtypes of dementia.

Recently, first clinical evaluation studies have become available for non-commercial, research-level and open-source tools for brain morphometry. In small case-control studies focusing on hippocampal sclerosis in temporal lobe epilepsy, Goodkin et al. (2021) and Rebsamen et al. (2023c) have compared expert ratings without and with availability of quantitative reports (QReports). Both found that with QReports available the accuracy and rater confidence for presence of hippocampal sclerosis increased, whereas disagreement among experts reduced. An obvious next step of our research will be to conduct similar studies with our open-source tool ScanOMetrics. Depending on the clinical question and suspected disease, different quantitative findings are in the center of the user’s interest. To ease ScanOMetrics usage, we will develop a graphical user interface (GUI) and design disease specific QReports.

## Data availability

The openly available OASIS3 dataset (2’842 longitudinal MRIs from 1’379 participants, many with re-scans, some with same-session re-scans, normal aging and Alzheimer’s disease) was downloaded from https://www.oasis-brains.org/#data

## Code availability

All software tools used in this paper are open source. The Python3 implementation of ScanOMetrics is available at https://github.com/SCAN-NRAD/ScanOMetrics. A code description is given in the Supplementary Materials and a more detailed documentation with tutorial is available at https://readthedocs.scanometrics.io. FreeSurfer can be downloaded from https://surfer.nmr.mgh.harvard.edu/ and DL+DiReCT is available at https://github.com/SCAN-NRAD/DL-DiReCT.

## Author contributions

Conceptualization: DR, CR; Methodology: DR, MR, PR, CR; Software: DR, MR, CR; Formal analysis: DR, CR; Resources: RW; Data Curation: MR; Writing original draft: DR, CR; Writing review and editing: all co-authors; Supervision: RM, CR; Project administration: RW, CR; Funding acquisition: RW, CR

## Acknowledgements

This work was supported by Swiss National Science Foundation (SNSF) under Grant/Award Numbers: 204593 (ScanOMetrics) and CRSII5_180365 (The Swiss-First Study).

