## Supplementary for "Cortical thickness and grey-matter volume anomaly detection in individual MRI scans: Comparison of two methods"

### Supplementary figures

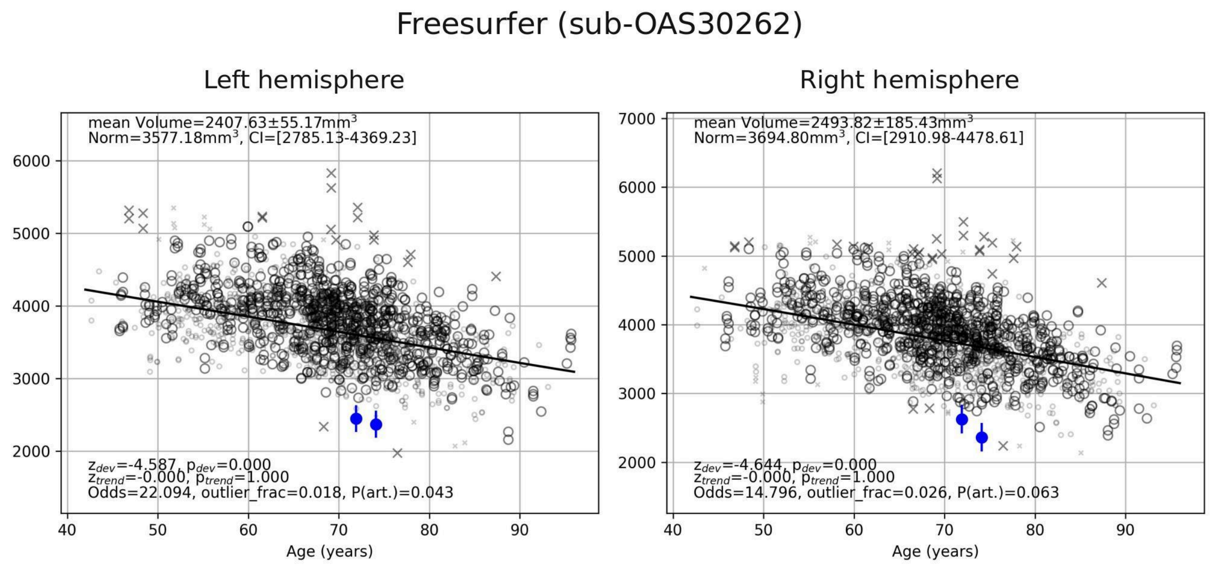


Supplementary Figure S1: Normalised volumes of the hippocampus of subject OAS30262 estimated by Freesurfer. Results are similar to the evaluation made using DL+DiReCT derived volumes (see main Figure 4). Even though the reference trajectory is steeper when considering Freesurfer volumes, the z-score for the subject is higher when using DL+DiReCT, and the decrease in volume between the two scans is higher when using DL+DiReCT.

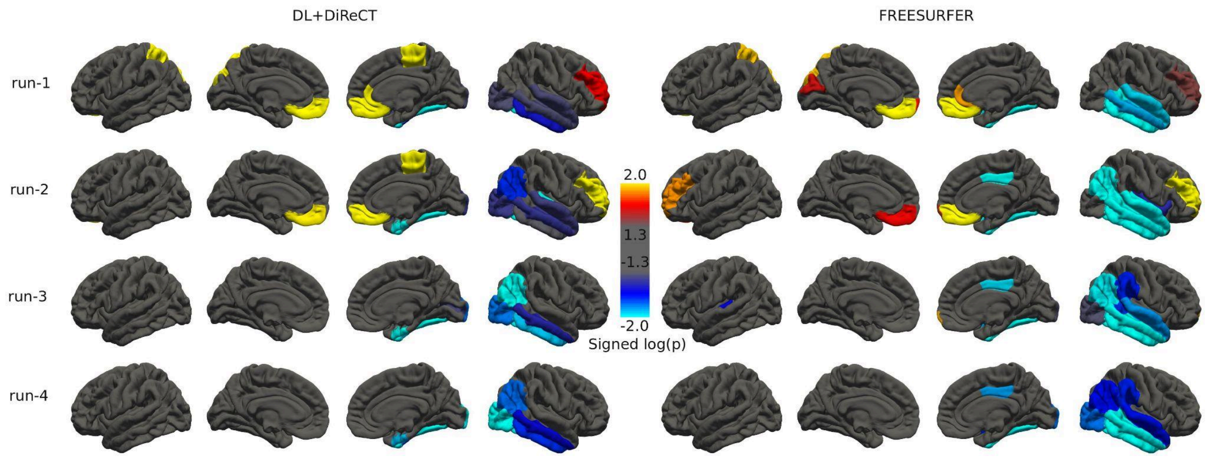

Supplementary Figure S2: log10(p) maps for the subject with CDR>=2.0 and the highest number of scans (OAS30902). Visual inspection of the MRI shows that runs 1 and 2 are blurred, likely due to patient motion during the scan, which could explain the decision to acquire runs 3 and 4, which have better image quality and higher grey-white contrast. We attribute significant CTh increase in the first two scans to the blurring artefacts. Thinning of the cortex in the right temporal lobe is likely a true finding, as it appears to be reproducible over all scans (including those with high image quality), and robust to subject motion artefacts.

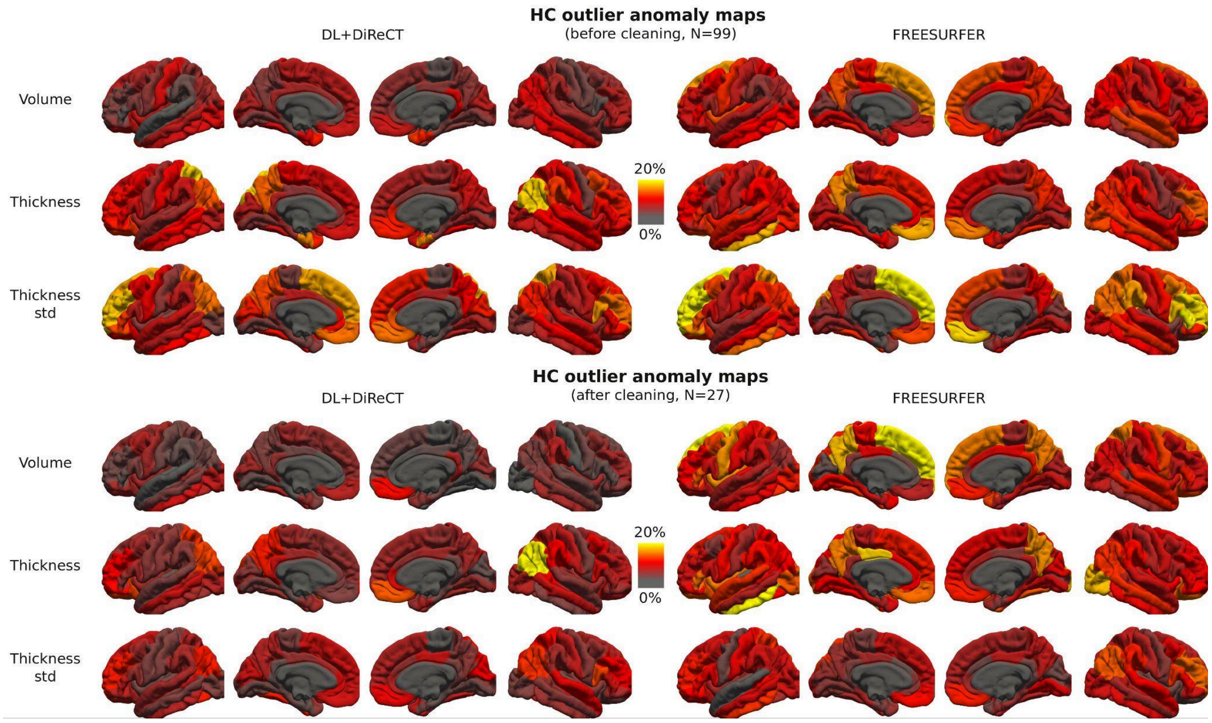


Supplementary Figure S3: Maps of the spatial distribution of abnormal metrics (GMV, mean and standard deviation of CTh) for outliers in the HC dataset, before and after our cleaning procedure. Outliers correspond to scans with more than 5% abnormal regions, using metrics derived from either DL+DiReCT or Freesurfer.

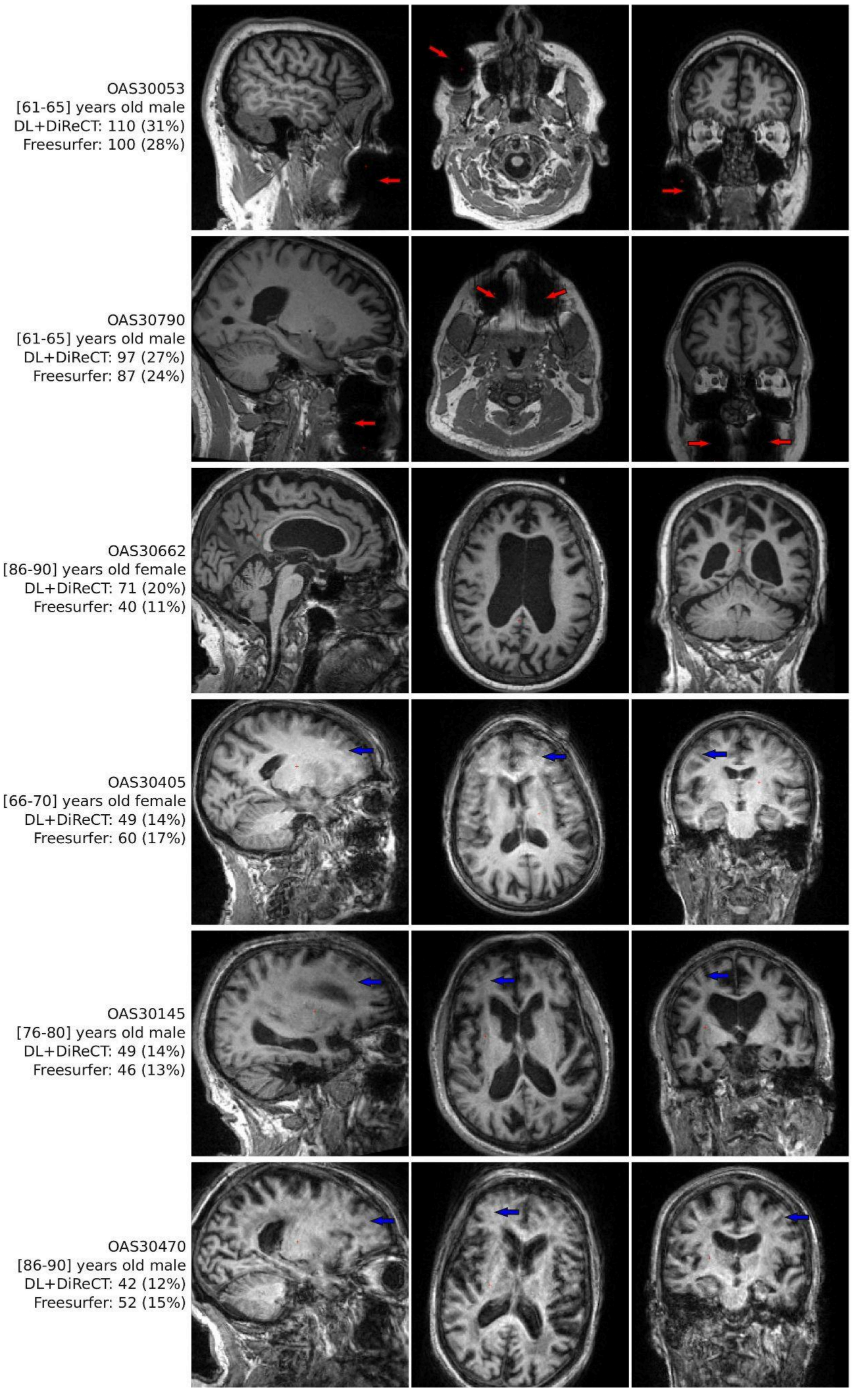


Supplementary Figure S4: Visual inspection of the outlier scans in the HC dataset that had the highest number of abnormal regions before cleaning. Selected scans show susceptibility artefacts (red arrows, likely due to dental implants) and blurring/ringing (blue arrows, likely due to subject motion during the scan). Subjects OAS30053 and OAS30790 have another scan with the same susceptibility artefact, which had similar numbers of abnormal regions despite the apparent good image quality inside the brain. In subject OAS30662 image quality is high, but the subject has enlarged CSF space suggestive of atrophy, thinned corpus callosum, and prominent lateral ventricles, larger than expected at this age (and was confirmed by ScanOMetrics labelling about 20% of metrics as abnormal). Number of abnormal regions detected by ScanOMetrics using either DL+DiReCT or Freesurfer are reported on the left of each scan, with percentage over the number of metrics in parenthesis.

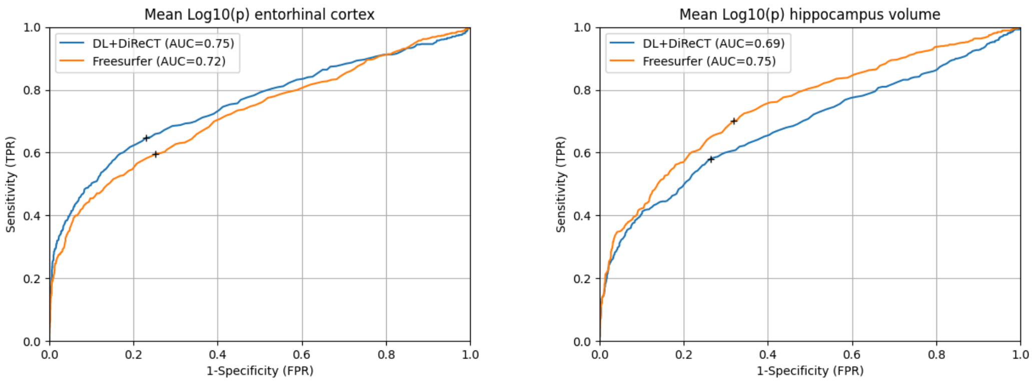


Supplementary Figure S5: ROC curves for averaged log10(p) values of entorhinal thickness (left plot) and hippocampal volume (right plot). DL+DiReCT and Freesurfer True Positive rate (TPR) are plotted against the False Positive Rate (FPR) for different thresholds of log10(p) values. Black crosses show the threshold for which the sensitivity and specificity were the closest to the top-left corner. For the entorhinal cortex, the threshold was -0.26 for DL+DiReCT (FPR=0.23, TPR=0.65) and -0.24 for Freesurfer (FPR=0.25, TPR=0.60). For the hippocampal volume, the threshold was -0.25 for DL+DiReCT (FPR=0.26, TPR=0.58), and -0.18 for Freesurfer (FPR=0.31 and TPR=0.69).

### Supplementary materials

Detailed documentation regarding ScanOMetrics can be found on https://scanometrics.readthedocs.io. The following is a summary of the different steps involved in two use-cases referred to in the main manuscript: the creation of the two normative models, as well as the evaluation of scans with CDR >= 1.

***Creating normative models***

1) *File structure:* ScanOMetrics expects study files to be organised according to BIDS recommendations (https://bids-specification.readthedocs.io/en/stable).

    a) Study data should be stored in a study folder. Each participant should have a single folder with its participant ID (i.e. sub-XXX) and the participants.tsv file should list all participants exactly once. We recommend using a session folder (i.e. ses-XXX) for each session, in particular when new scans might be expected.

    b) Covariates: Immutable covariates should be included in the participants.tsv file, while covariates that might change between scans (e.g. age, sequence, scanner) should be included in the participant's sessions.tsv file.

2) *Study initialization and subject loading:* A ScanOMetrics object is initiated by specifying the path to the BIDS folder. A "cov2float" dictionary can be specified to map categorical covariates to numerical values (e.g. the 'sex' covariate value 'f' could be mapped to "1" and "m" to "0"). When loading subjects, a dictionary with all sessions and scans is created, along with covariate values, following the content of participants and session tsv files. Subjects, sessions and specific scans can be included/excluded by listing them in the "subjects_include", "subjects_exclude", "sub_ses_include", "sub_ses_exclude", "sub_ses_acq_include" or "sub_ses_acq_exclude" arguments from the load_subjects() method.

3) *MRI processing*: ScanOMetrics' MRI processing modules currently include Freesurfer and DL+DiReCT. Atlas options include the Desikan-Killiany or Destrieux atlas. DL+DiReCT can also process contrast enhanced scans. In the current study, scans were processed with Freesurfer and DL+DiReCT, using default parameters (both based on the Desikan-Killiany atlas and no contrast enhancement for DL+DiReCT). ScanOMetrics calls the processing pipeline with parameters set by the user, and processes each scan loaded from the dataset. After processing, SBA values are saved in the BIDS "derivatives" directory, in a folder named after the processing tool, and a sub-folder for each scan. Scan folder names are flattened in terms of subject, session and acquisition IDs (one folder per scan).

4) *Computation of normative model*: takes as input the raw SBA values computed by an MRI processing software (i.e. FreeSurfer or DL+DiReCT) and fits the normative model of interest (e.g. a polynomial model). This step needs to be done only once per study and can be loaded for normative evaluation of new subjects, see section on evaluation of new scans. The parameters of the MRI processing as well as the specification and demographics of the normative dataset are saved within the same pkl file.

a) *Metric loading and complementary metric computation:* SBA values computed in the previous step are loaded into a single matrix (one column per metric and one row per scan). Loading will depend on the pipeline and available files. For example, in the current study, DL+DiReCT computes volume, thickness and standard deviation of the thickness, whereas FreeSurfer also computes additional metrics like WM/GM contrast, curvatures, folding index, etc… Metrics to be loaded can be specified through the "metrics_include" argument in the "load_proc_metrics()" method. ScanOMetrics computes a few additional metrics like the symmetry index for metrics defined bilaterally, as well as lobe metrics by aggregating corresponding regions of interest. Brain size normalisation with respect to the ratio between each subject's total intra-cranial volume (ICV) and the group's average ICV is also performed. Exponents are taken according to the metric's dimensionality (exponents of 1, 2/3, and 1/3 are taken for volumes, areas, and lengths, respectively). In the scope of the current study, this translates into 239 additional 'normalized' values.

b) *Initial outlier detection:* for each metric, SBA values deviating more than 1.5 inter-quartile ranges (IQR) from the upper or lower quartile of the distribution are labelled as outliers, and excluded from the subsequent steps. The IQR is computed for each scan separately, by considering the distribution of age-matched SBA values (scans with age within 10% of the age of the scan of interest).

c) *Initial polynomial fit:* to mitigate effects of potentially non-uniform sampling of age in the normative dataset, ScanOMetrics subsamples the distribution N times. An age histogram is computed using 10 bins by default. The least filled bin defines the number n of scans to be randomly drawn from all bins during the subsampling procedure, finally leading to 10*n scans in each of the N subsamples. The age distribution of each subsample is uniform to a good approximation, with the caveat that the set of scans in the smallest bin is present in all the N subsamples. ScanOMetrics fits a low order polynomial age model to each metric separately for all N subsamples. Starting at zero, the polynomial degree is gradually increased until no further statistical improvement of the fit is observed (nested F-tests). The maximum degree was set to 20 to avoid uncontrolled overfitting. This procedure leads to N polynomial fits for each metric, with parameters depending on details of the randomly subsampled set. Fit residues are computed by taking the difference between the measured value and the average of the predictions by the N polynomial models.

d) *Uncertainty estimation:* an upper bound for measurement uncertainty was estimated based on the variance of metrics across scans repeated within 10% of the subject's age, discarding outliers found in the previous step.

e) *Final outlier detection based on residuals:* a second outlier detection loop similar to step c) discards scans with residuals farther than 1.5 IQRs from the upper or lower quartile.

f) *Final polynomial fit:* a second polynomial fit is performed for each metric separately, as in step c), this time also discarding scans labelled as outliers in step e).

The degrees of all metrics and uniform subsamplings can be saved in a pkl file, along with all information required for further scan evaluation and plotting (processing pipeline parameters, measured metrics and fit residues, uncertainty estimates, outlier list, covariates, cov2float dictionary, etc…), using the "save_normative_model()" method. We recommend saving pkl files to a dedicated folder in the ScanOMetrics python package ("scanometrics/resources/normative_models"). This allows ScanOMetrics to list both distributed and user specific normative models, through the "list_normative_models()" method.

***Evaluating new scans***

1) *File structure:* ScanOMetrics expects study files to be organised according to BIDS recommendations (https://bids-specification.readthedocs.io/en/stable).

    a) Study data should be stored in a study folder. Each participant should have a single folder with its participant ID (i.e. sub-XXX) and the participants.tsv file should list all participants exactly once. We recommend using a session folder (i.e. ses-XXX) for each session, in particular when new scans might be expected.

    b) *Covariates:* Immutable covariates should be included in the participants.tsv file, while covariates that might change between scans (e.g. age, sequence, scanner) should be included in the participant's sessions.tsv file.

2) *Study initialization:* A ScanOMetrics object is initiated by specifying the path to the BIDS folder.

3) *Loading of pretrained normative models:* ScanOMetrics is distributed with a selection of pre-trained normative models, which can be listed through the list_normative_models() function. This function lists pkl files available in the dedicated ScanOMetrics folder ("scanometrics/resources/normative_models"), which can be user trained models or models distributed with ScanOMetrics. Model loading is achieved by running the load_normative_model() function. If available in the ScanOMetrics folder, normative models can be loaded by giving the filename with or without the pkl extension. Other models can be loaded by giving the full path to the pkl file, including file name and pkl extension. To ensure that new scans are processed identically to the loaded normative model, the MRI processing parameters and the dictionary to convert categorical covariates to values ("cov2float") are loaded from the normative model's pkl file. In the context of the current study, the normative models are "Polynomial_dldirect_cleanOASIS3" and "Polynomial_freesurfer_cleanOASIS3", which are polynomial fits to SBA values estimated using either FreeSurfer or DL+DiReCT, on the OASIS3 scans with CDR=0 after removing outlier scans (see main text for details on the additional cleaning procedures).

4) *Subject loading*: a subject dictionary with all sessions and scans is created by calling the "load_subjects()" method. The method loads subjects according to session and acquisition labels, along with covariate values, following the content of participants and session tsv files. Subjects, sessions and specific scans can be included or excluded (see normative modelling pipeline).

5) *New subject processing:* the scans from a new subject can be processed using the same processing tool (i.e. either FreeSurfer or DL+DiReCT as stored in the specification of the normative model, see step 3) of the normative modelling section) and its SBA metrics are saved to a table. All processing outputs are saved into the BIDS "derivatives" folder, in a folder dedicated to each processing pipeline (e.g. "freesurfer" or "dldirect") and in one subfolder per scan (flattened structure). SBA metrics are then loaded and complementary metrics computed, as done for the normative dataset. Since ICV values are available from the normative dataset, normalised metrics are computed by taking average values from the normative dataset as reference.

6) *New subject evaluation:* all scans of a new subject can be evaluated against the normative model using the "evaluate_singleSubject_allSes()" method, and specifying the subject ID to be evaluated. The "matching_covariates" argument allows specifying categorical covariates used to determine matching scans in the normative dataset to evaluate against (default set to 'sex', 'sequence' and 'scanner'). For each metric, ScanOMetrics computes the residue by taking the difference between the measured value in the new scan and the average of N values expected from the
